# Tuberculosis services delivery challenges and their mitigations during the COVID-19 pandemic in Tanzania: A qualitative study

**DOI:** 10.1101/2025.03.19.25324251

**Authors:** Doreen Pamba, Erica Sanga, Wiston William, Happiness Mvungi, Hamimu Omary, Theresia Setebe, Willyhelmina Olomi, Chacha Mangu, Issa Sabi, Robert Balama, Emmanuel Matechi, Riziki Kisonga, Allan Tarimo, Nyanda Elias Ntinginya, Lucas Maganga

## Abstract

**Objective:** To describe challenges posed by COVID-19 on TB commodity supply, care cascade, active case finding, and responses taken by healthcare workers (HCWs) and community health workers (CHWs) during the first year of the pandemic (March 2020 to February 2021).

**Design:** A qualitative descriptive study involving 25 in-depth interviews and 10 focus group discussions conducted in July 2022.

**Setting:** 37 TB treatment facilities were purposively selected from seven regions due to high TB case notifications in 2019 and their provision of TB and COVID-19 services during the first year of the pandemic (March 2020 to February 2021).

**Participants:** Purposive selection of 58 HCWs and 55 CHWs who provided TB services in the first year of the COVID-19 pandemic.

**Results:** HCWs reported unusual stockouts and delayed receipt of GeneXpert cartridges and sputum containers. TB services faced a decline in client attendance, as clients were hesitant to undergo TB screening, sputum sample collection, and contact tracing due to fear of contracting or being diagnosed with COVID-19 and subsequently being quarantined. To mitigate these challenges, HCWs used alternative containers for sputum sample collection, optimized GeneXpert cartridges use by prioritizing GeneXpert testing for TB risk groups, and diagnosed TB by microscopy, chest X-ray, and sputum pooling method. Moreover, they extended drug refill schedules to minimize the risk of contracting COVID-19 in clinics. CHWs used mobile communication for client tracing and focused household visits on TB risk groups.

**Conclusion:** COVID-19 disrupted TB commodity availability and TB treatment-seeking behavior. Adaptations like multi-month drug refills and optimized GeneXpert use, supported the TB healthcare system’s resilience. While these adaptations offer valuable insights for strengthening TB service delivery, their effectiveness and sustainability require further evaluation. Thus, prospective studies could clarify their long-term impact. National Tuberculosis Programs could consider adapting these practices post-pandemic, with appropriate modifications to suit different contexts.

**STRENGTHS AND LIMITATIONS OF THIS STUDY:**

- The study provides insights into healthcare providers’ TB management experiences during the first year of COVID-19.
- Purposive sampling provided insights into TB service delivery across diverse healthcare cadres and facilities in high-COVID-19 regions.
- Triangulation of data collection methods and researchers enhanced credibility by ensuring data consistency and reducing potential bias.
- Retrospective data collection may have introduced recall bias but mitigated through data triangulation.
- Focusing on high-COVID-19 regions may limit the transferability of findings to less COVID-19 affected areas with different TB service challenges.

## INTRODUCTION

The coronavirus disease (COVID-19) ranks first as the leading cause of death from a single infectious agent worldwide, surpassing deaths from tuberculosis (TB) that follow second^1^. As of January 1, 2023, the global mortality cost from COVID-19 was $29.4 trillion^2^. The WHO reported over 775 million confirmed cases of COVID-19 and more than 7 million deaths globally on 21^st^ July 2024^3^. In Tanzania, the burden of COVID-19 cases varied across regions^4–6^. Until 23^rd^ December 2022, the Tanzania Ministry of Health announced 36,969 confirmed COVID-19 cases and 808 deaths from COVID-19^7^.

The pandemic has caused significant disruptions in the delivery of healthcare services worldwide, including those for TB that vary in magnitude and duration^8–11^. Globally, 51% of the 98 countries that responded to a WHO survey reported disruptions in TB diagnosis and treatment during the first year of the COVID-19 pandemic, ranging from 5% to >50%^12^. The disruptions were also reported to have plateaued in the second year of the pandemic, with a follow-up WHO survey that reported disruptions in 49% of 90 countries^13^. A decrease of 18% in global TB case notifications was observed in 2020 compared with 2019^14^.

Tanzania is among the 30 high TB/HIV burden countries in the world^1,15^. The country was reported to have a minimal increase in TB incidence during and after the pandemic, whereby an increase of 16.3% in all forms of TB was observed between 2021 and 2022^16^. Moreover, during the COVID-19 pandemic, the WHO categorized it as among the six high TB burden countries that experienced limited disruptions to TB detection between 2020 and 2022^17^. The country notified 85,120, 86,661, and 100,747 all forms of TB cases in 2020, 2021, and 2022, respectively, compared to 81,492 cases in 2019^16^. This shows steady TB case notification rates amidst the pandemic.

Although evidence exists of the impact of COVID-19 on varied aspects of TB management among several countries^18^, available studies in Tanzania describe the effects of the pandemic on other health aspects, namely HIV, reproductive health, and mental health^18–20^, and not on TB. Furthermore, the WHO recommends that countries research the unknowns of the COVID-19 pandemic^21^. Understanding TB service delivery in Tanzania during COVID-19 period is salient for improving post-pandemic TB management and future pandemic preparedness. This qualitative study is part of a mixed methods study (S1 File) where the quantitative component retrospectively assessed the pandemic’s impact on TB services delivery by comparing two equivalent periods: pre-COVID-19 (March 2019 to February 2020) and during COVID-19 (March 2020 to February 2021). This study, therefore, describes challenges posed by COVID-19 on TB commodities supply, TB care cascade, active case finding, and responses taken by healthcare workers (HCWs) and community health workers (CHWs) during the first year of the pandemic (March 2020 to February 2021).

## METHODS

### Study design

We used a qualitative description approach to describe TB services during the first year of the COVID-19 pandemic in Tanzania. This period coincided with the first and second COVID-19 waves, where healthcare providers would have implemented varied coping mechanisms offering valuable lessons for future pandemic preparedness. The study design is suitable for providing an in-depth understanding of a phenomenon with interpretation limited to a literal description of the data collected^22,23^.

### Study setting

The study was conducted across seven regions in Tanzania: Dar es Salaam, Arusha, Kilimanjaro, Mwanza, Mbeya, Dodoma, and Kigoma. The regions were purposively selected due to high COVID-19 cases during the pandemic’s first year, and high cross-border socio-economic activities in Kigoma, Kilimanjaro, and Mbeya hypothesized to facilitate easy spread of SARS-CoV-2. Furthermore, 37 TB treatment facilities were purposely selected within the regions for their high TB case notifications during the pre-COVID-19 period (March 2019 to February 2020) and operated as COVID-19 centers during the pandemic’s first year while continuing TB services provision.

### Study sampling and population

We purposively sampled healthcare providers across the 37 TB treatment facilities including; doctors, nurses administering directly observed therapy (DOT nurses), regional TB and Leprosy coordinators (RTLCs), district TB and Leprosy coordinators (DTLCs), district TB HIV officers (DTHOs), and CHWs assisting active case finding (ACF) in the same treatment facilities. After obtaining regional permissions, we confirmed participation with RTLCs and DTLCs by phone. With approval from Medical Officers In-charge (MoIs), we approached doctors, DOT nurses, and CHWs in the respective treatment facilities, all of whom accepted participation and scheduled interviews at their convenience. Participants had to provide informed consent and have worked in the respective TB treatment facilities during the first year of COVID-19; those on extended leave or not directly involved in TB care during that time were excluded.

### Data collection

Data were collected in July 2022 by trained Social Scientists with expertise in qualitative research (including DP, ES, and TS) and Public Health experts (HM and LM) experienced in mixed methods studies. The team, comprising two males and four females with Bachelor degrees in Sociology or Master in Public Health, were Research Scientists experienced in TB program evaluation but not directly involved in TB service provision, thus minimizing social desirability bias.

Semi-structured in-depth interviews (IDIs) and focus group discussions (FGDs) were audio-recorded in Kiswahili for approximately 45 to 90 minutes at respective TB treatment facilities in a room that guaranteed privacy. IDIs were conducted with nine RTLCs, four DTLCs, three DTHOs, and nine CHWs. Five FGDs were conducted in five regions with 42 HCWs (facility doctors, DOT nurses, and laboratory technicians), each comprising 6-12 participants. Additional five FGDs of 6-12 participants were conducted with 46 CHWs. Code saturation was reached at the 16^th^ HCW IDI transcript following a team agreement (DP, ES, HM, LM, and HK) during the iterative coding process. Discussions focused on COVID-19 risk perceptions, prevention strategies, challenges in TB commodities availability and TB care cascade, and measures taken (S2 File, S3 File, S4 File, and S5 File).

### Data analysis

The interviewers transcribed the audio recordings verbatim and translated them from Kiswahili to English. To ensure the accuracy of verbatim transcription, DP and ES independently reviewed all transcripts against the audio recordings and made corrections accordingly, thereby facilitating data familiarization.

The transcripts were analyzed using thematic framework approach. DP developed initial coding matrices using structured coding based on research questions and interview guides, coding the first two sets of IDIs and FGDs transcripts for HCWs and CHWs. These were later reviewed by ES, LM, HM, HK, and TM, thus, enabling the team’s familiarization with the data. During team review, new codes and categories were developed, collapsed, renamed, or reordered, thus refining the codebooks. Upon team agreement on the final codebooks and thematic frameworks, DP, ES, HM, LM, and HK independently coded the remaining transcripts. Although 25 IDIs were held with HCWs, during coding, nine IDI transcripts with HCWs were identified as lacking richness in addressing the research questions largely due to brief participant responses despite probing. The team agreed that the transcripts lacked new insights thus, excluded in subsequent analysis to maintain analytical depth. To strengthen the robustness of our findings, we cross-checked code saturation against FGD transcripts. The coded data were then grouped into categories, and themes were developed through iterative team discussions. The team worked collaboratively to ensure that the themes captured the depth and diversity of participants’ experiences and reflected a logical flow of ideas. Relevant quotations were selected to illustrate the themes comprehensively.

To ensure trustworthy findings, we triangulated methods and researchers by using IDIs and FGDs administered by different interviewers and involved multiple investigators in data analysis. This generated diverse perspectives, enhancing the study findings’ credibility. We provided a thick description of participant characteristics and the conditions under which TB services were delivered during the COVID-19 pandemic, therefore enabling readers to determine the transferability of the findings to similar contexts or populations.

### Patient and public involvement

During the protocol development phase, we engaged eleven TB frontline HCWs from the seven regions and five National Tuberculosis and Leprosy Control Programme (NTLP) staff. Their input informed the development of data collection tools, logistics, and dissemination strategies.

### Ethical considerations

Ethical approval was sought from the Mbeya Medical Research Ethics Committee (MMREC *Ref. SZEC-2439/R.A/V.1/147a*). Research permission at health facilities was granted by the President’s Office, Regional Administration and Local Government (PORALG), Regional Administration, and facility MoIs. Written informed consent was obtained from participants, who were informed about the study’s purpose and their rights. We assigned unique codes to transcripts, removing identifying information for confidentiality. Data were securely stored and accessible only to authorized team members.

## RESULTS

The majority of the HCWs (23/58) worked at district hospitals and had experience providing TB services for about five years (Table 1).

**Table 1:**
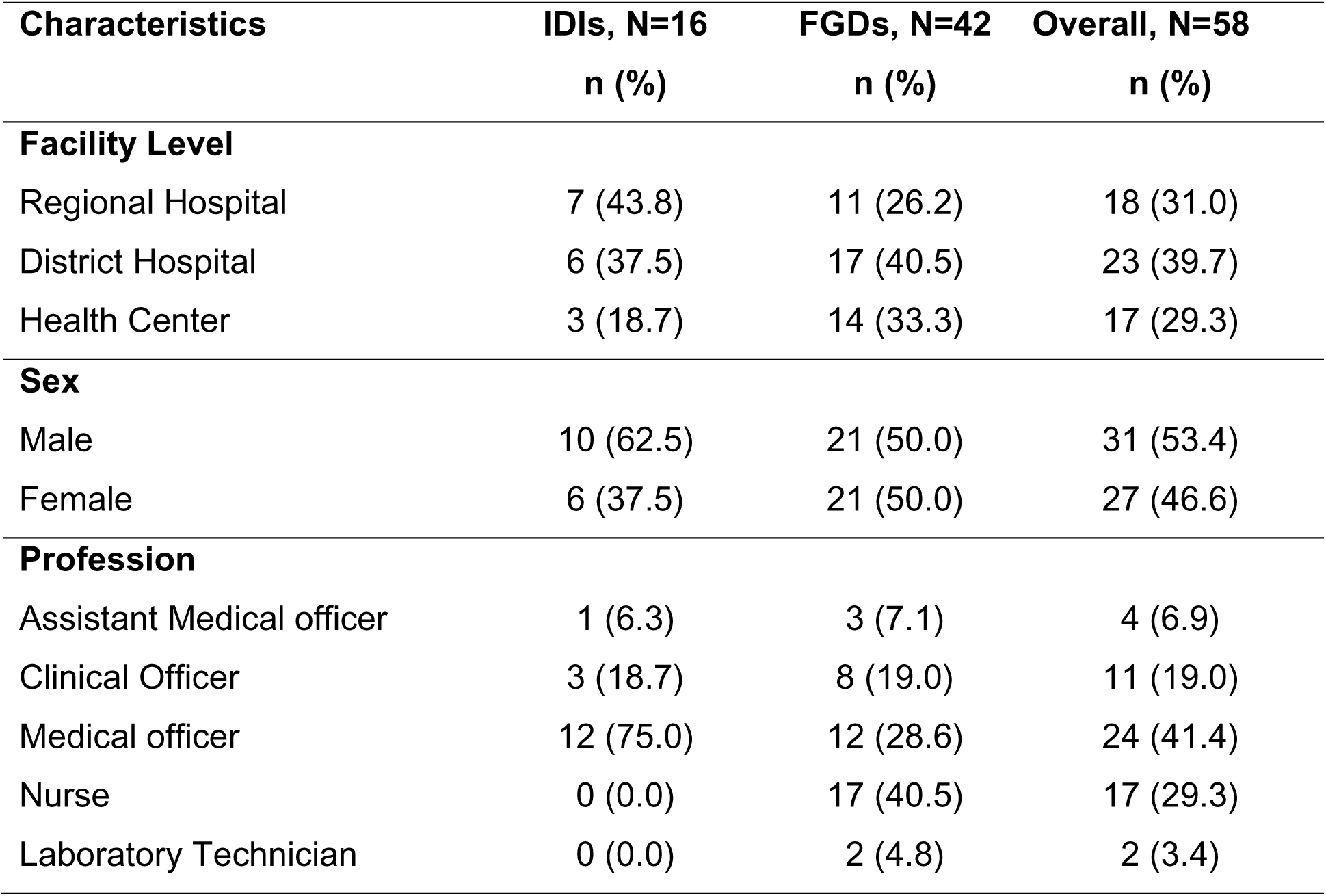
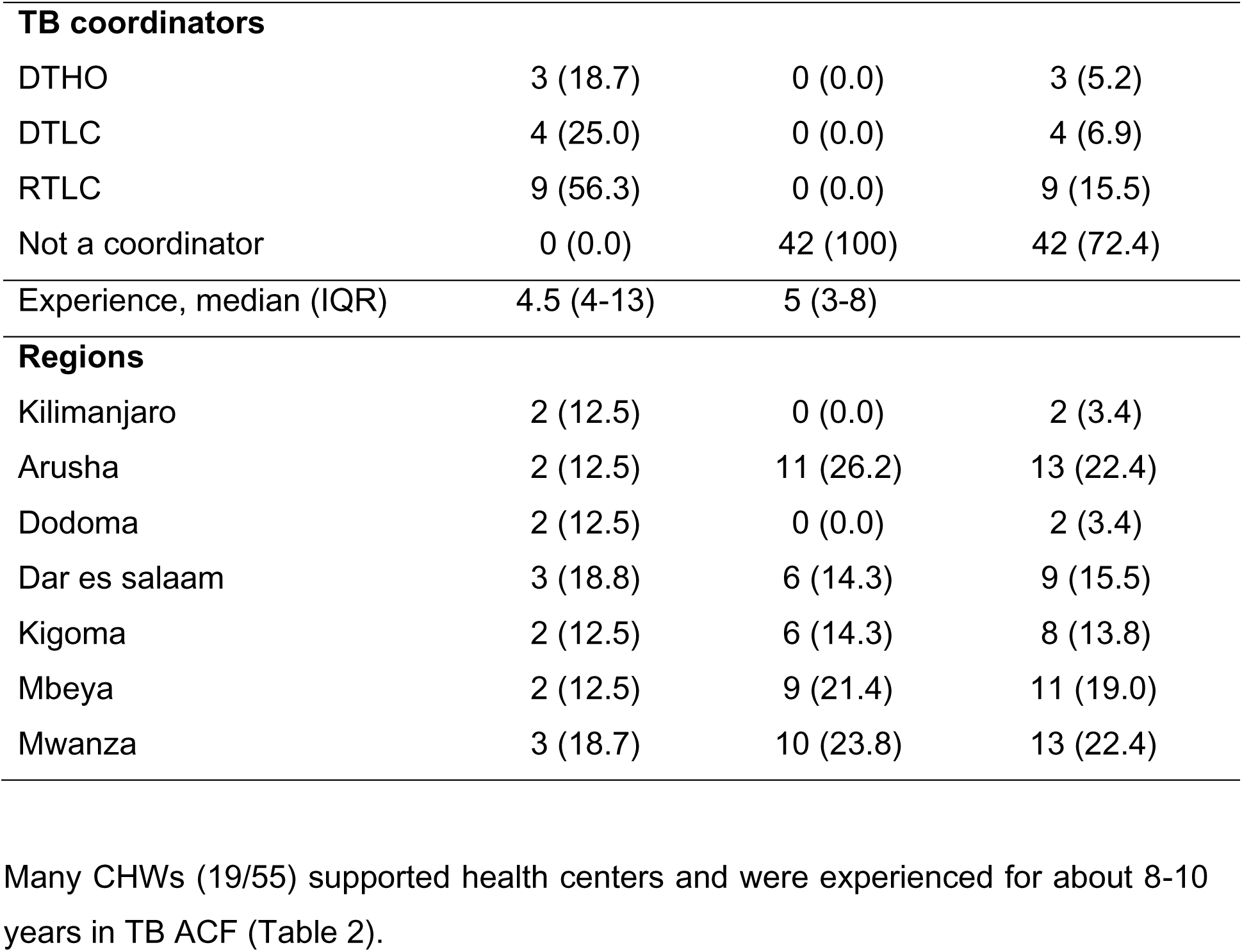
Characteristics of HCWs in IDIs and FGDs.

Many CHWs (19/55) supported health centers and were experienced for about 8-10 years in TB ACF (Table 2).

**Table 2:**
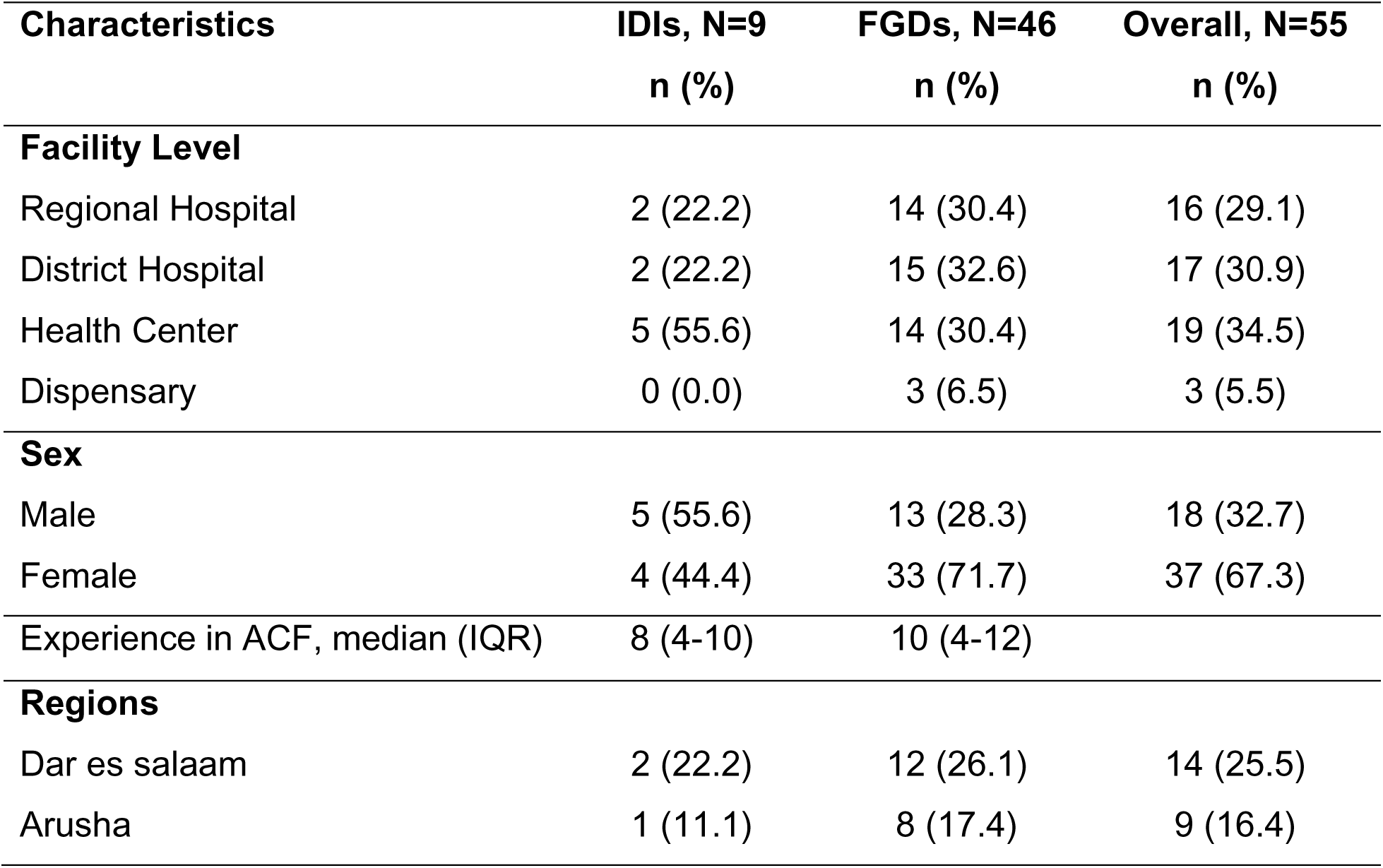
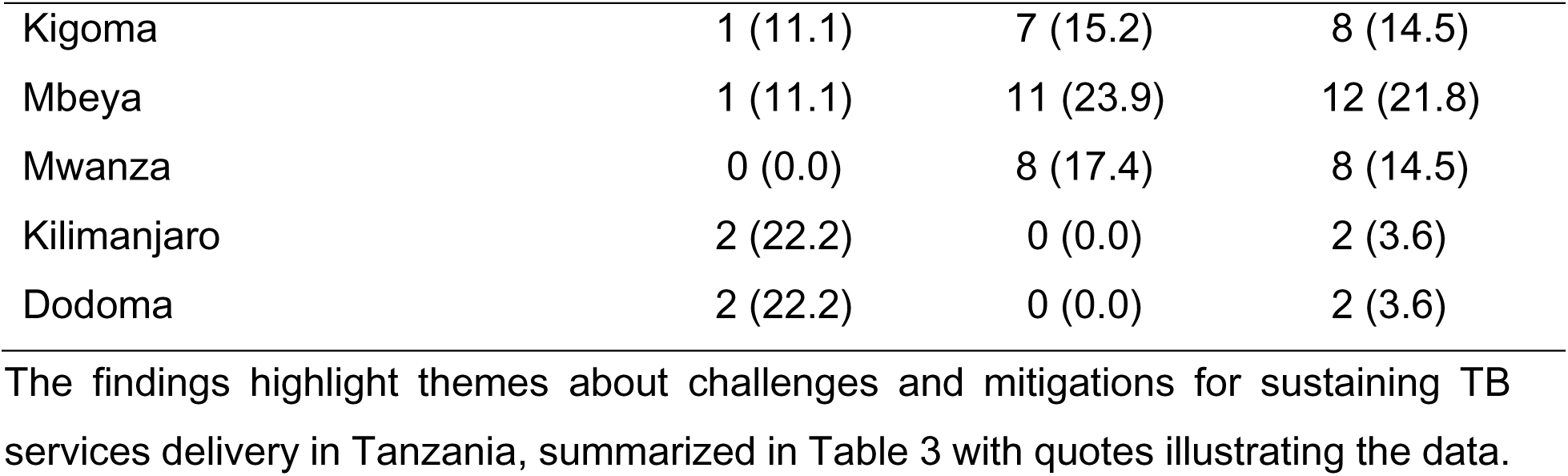
Characteristics of CHWs in IDIs and FGDs.

**Table 3:**
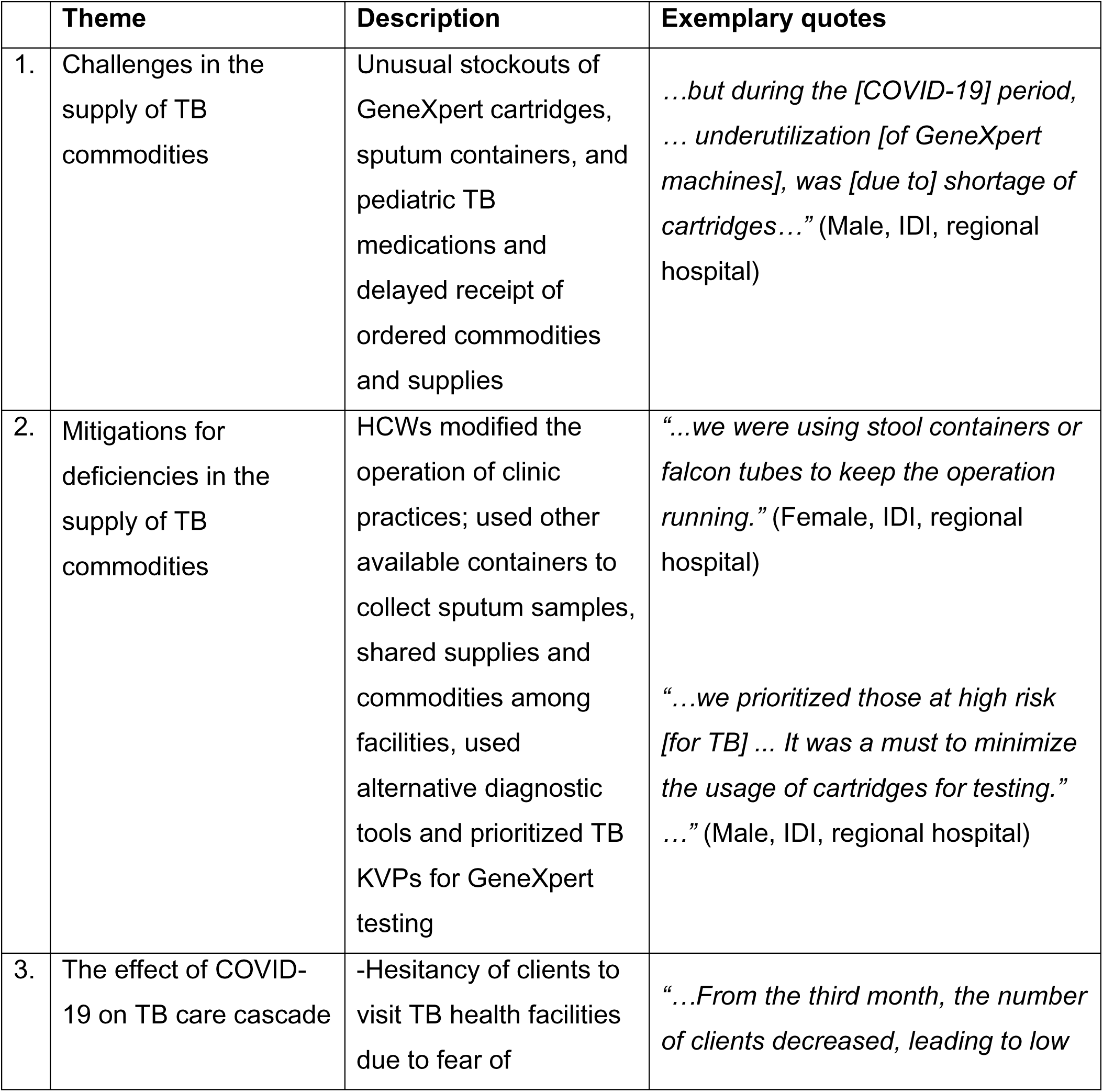
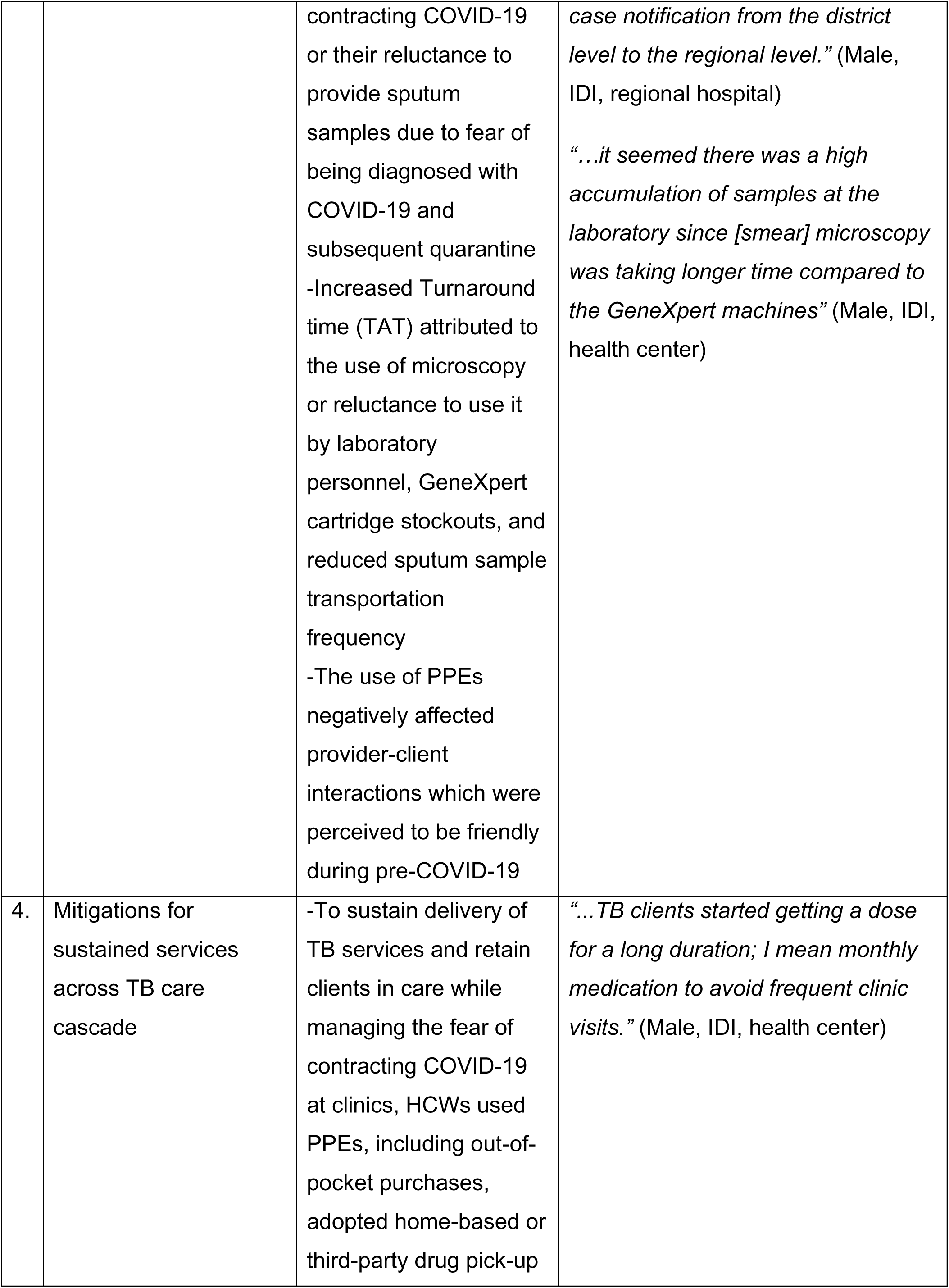
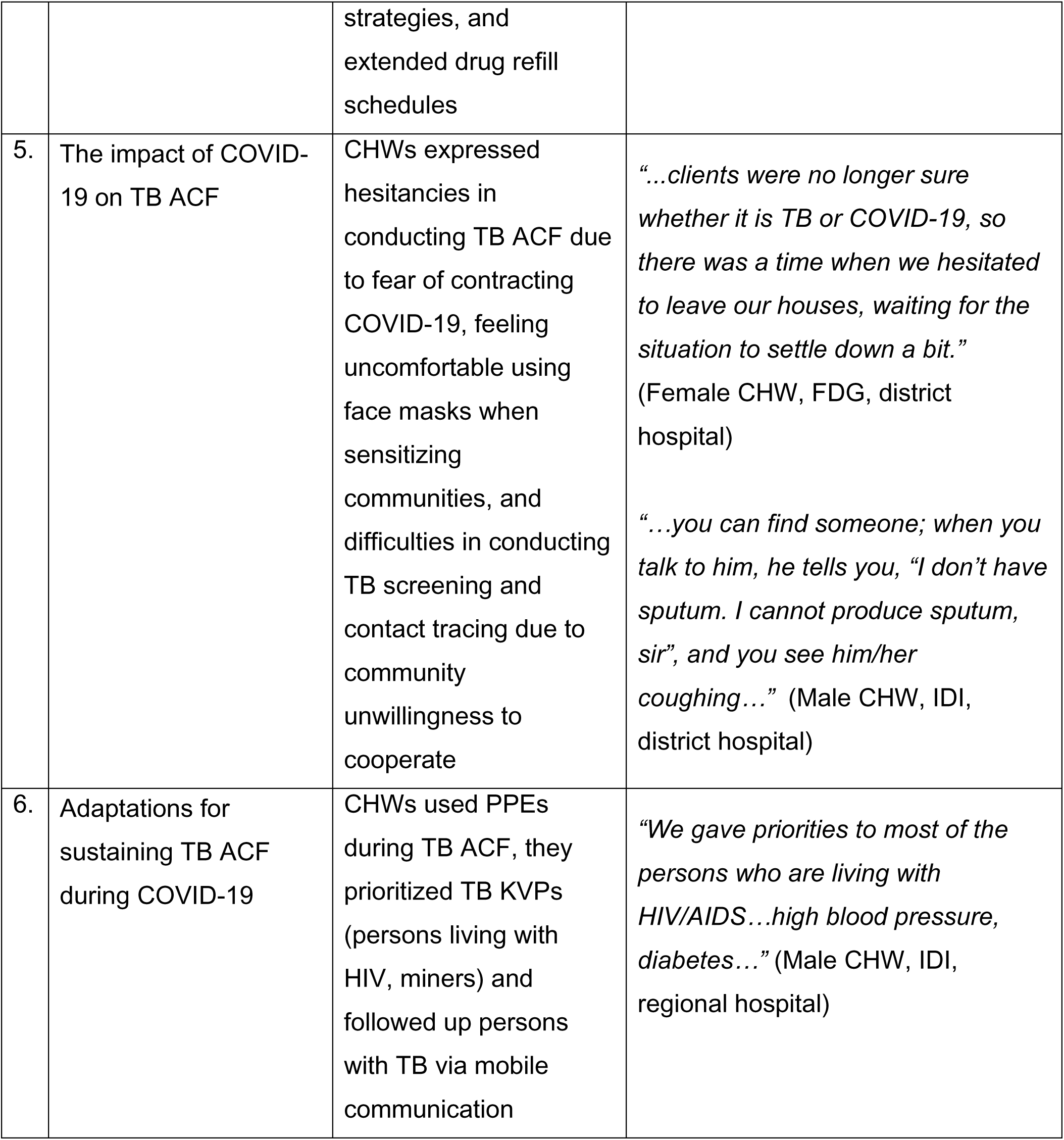
Summary of the key themes.

### Challenges in the supply of TB commodities

HCWs reported unusual stockouts and delays in receiving TB commodities ranging from three weeks to six months. Stockouts of GeneXpert cartridges, sputum containers, and N95 masks were encountered across all facility levels. One HCW attributed cartridge stockouts to the underutilization of GeneXpert machines during the pandemic:

> *“…back [in] 2018, 2019…the ministry was concerned about low utilization of GeneXpert machines. Although cartridges were available, they were not adequately used for testing…but during the [COVID-19] period… underutilization, was [due to] shortage of cartridges…”* (Male, IDI, regional hospital)

Furthermore, HCWs reported shortages in pediatric TB drugs and receipt of less quantity of commodities contrary to what was ordered from the Medical Stores Department:

> *“…TB drugs for the children, we were staying even more than a month without receiving the order.”* (Male, IDI, district hospital)
>
> *“…we received [commodities] depending on what was available. So, even the Medical Stores Department didn’t consider how much facility X or Y requested.”* (Male, IDI, health center)

### Mitigations for deficiencies in the supply of TB commodities

HCWs adapted clinical practices to mitigate TB commodity shortages by using alternative sputum sample collection containers and efficiently using available commodities by leveraging TB screening to increase the likelihood of detecting TB.

> *“…despite the shortage, we were using stool containers or falcon tubes to keep the operation running, we didn’t stop”* (Female, IDI, regional hospital).
>
> *“…we made sure we had a detailed history taking and minimized doubts, then those who were suspected [to have TB disease were] the ones who provided the sputum [sample] to be tested by the GeneXpert machine. So, when someone went to provide the sputum samples, we were almost sure of the [TB] symptoms.”* (Male, FGD, regional hospital)

HCWs in facilities with surplus TB commodities distributed them to facilities facing shortages:

> *“…when we were about to run out of stock, we would borrow from other facilities…For example, I am a DTLC at [facility X]; in case I face a shortage, I would call [district Y], and if they don’t have enough, I would call another district for help…”* (Male, IDI, district hospital)

HCWs used alternative diagnostic platforms namely; microscopy and sputum pooling methods, including prioritizing TB high-risk clients for GeneXpert testing:

> *“…[sputum pooling] was performed at [facility X] … to minimize the use of cartridges…four samples were mixed, when it reads negative, then all are negative, but when it reads positive, then they will start testing one by one.”* (Male, FGD, district hospital)
>
> *“…we prioritized those at high risk [for TB]… to minimize the use of cartridges… For example, those who were previously treated… those from mining areas…also MDR-TB contacts…”* (Male, IDI, regional hospital)

Furthermore, adult TB drugs were a substitute for pediatric drug shortages:

> *“…but there is a period that the children’s medicine became a challenge so, sometimes…we used the adult doses…”* (Female, FGD, district hospital)

### The effect of COVID-19 on TB care cascade

HCWs continued TB screening services with COVID-19 prevention measures, but some reported a decrease in client attendance. This was attributed to clients preferring traditional medicine, believing their TB symptoms were mild, opting for home self-treatment, fearing COVID-19 infection or diagnosis with subsequent quarantine, and fearing unwillingly COVID-19 vaccination.

> *“…we started to find the clients, calling the family members, and we found out that the clients were already sent to traditional healers.”* (Male, IDI, district hospital)
>
> *“…the challenge was to counsel the client until he/she agrees to provide a [sputum] sample… because if you found out the client was tested here [at this facility] and was diagnosed with COVID-19 [and] he/she is at home, the ambulance comes… so the whole street sees that someonès house has COVID-19… many of them were hiding…”* (Female, IDI, regional hospital)

Few HCWs perceived the use of PPEs to have affected client-provider interactions from friendly to stigmatizing relationships:

> *“It was difficult, sometimes you can say that I am really stigmatizing this client when telling him/her to sit here and keep distance…there is no longer that friendship like it used to be before…”* (Female, IDI, district hospital)

TAT for sputum results varied. Some were delayed by four to seven days, while others were not. Delays were attributed to the use of microscopy or reluctance to use, GeneXpert cartridge stockouts, and reduced sputum sample transportation frequency:

> *“…during shortage [of cartridges], it was advised all clients suspected of TB to get tested through [smear] microscopy; however…[smear] microscopy was taking longer time compared to the GeneXpert machines”* (Male, IDI, health center)

Reports on TB diagnosis during COVID-19 varied. While some HCWs reported no changes, many reported low TB diagnosis rates due to stockouts of sputum containers and GeneXpert cartridges, decreased client attendance, reluctance to provide sputum samples, reduced sample transportation to testing facilities, and laboratory personnel’s fear of contracting COVID-19 from testing samples.

> *“…It was difficult to use smear [microscopy] in testing TB when you know exactly that COVID-19 is transmitted through contact. So, laboratory technicians stayed away from either TB clients or sputum…”* (Male, FGD, health center)

Some HCWs reported misdiagnosing TB clients as having COVID-19 and missed MDR-TB diagnosis due to difficulties in differentiating COVID-19 from TB during the early pandemic months:

> *“…there was a client… diagnosed with COVID-19, and admitted to the ward where he continued to receive COVID-19 treatment. He was not improving, but after five days, another doctor came and said that the client should also do a TB test; after testing, he was found to have MDR-TB.”* (Male, IDI, district hospital)

### Mitigations for sustained services across TB care cascade

HCWs took various precautions in clinic and community settings by wearing masks and gloves, using sanitizers, observing social distancing, washing hands with running water and soap, and getting vaccinated against COVID-19.

> *“…we got PPEs from different donors, including the Ministry of Health, who educated us about COVID-19. Different donors funded a lot of things, including sanitizers, soap…We had THPS, AMREF, and others.”* (Male, FGD, district hospital)

Nevertheless, there were stockouts of PPEs, leading HCWs and CHWs to purchase fabric masks out-of-pocket and to wear a single mask per day:

> *“…when we run out of masks, we were forced to use them without adhering to the required instructions…[although] we were instructed to change them after some hours, due to shortage, we were forced to wear them the whole day.”* (Male, IDI, district hospital)

HCWs provided drugs at clients’ homes, requested client relatives to support drug refills, and provided drugs at the facility gates to reduce COVID-19 risk for TB clients:

> *“…we were worried of losing clients, so sometimes, we allowed them to come at the gate and call us whereby we give them their medication [there]…”* (Female, FGD, district hospital)

Furthermore, all HCWs reported extending drug refill schedules to either two weekly or monthly, as recommended by the National Tuberculosis and Leprosy Programme and regional health management teams:

> *“…TB clients started getting a dose for a long duration; I mean monthly medication to avoid frequent clinic visits. So those who were in the intensive phase, we gave them a two-week dose instead of a week dose, and those in the continuation phase received a monthly dose.”* (Male, IDI, health center)

### The impact of COVID-19 on TB ACF

Mixed reports emerged on COVID-19’s impact on TB ACF, with some communities unaffected. CHWs reported barriers to TB screening and hesitations due to fear of contracting COVID-19:

> *“…clients were no longer sure whether it is TB or COVID-19, so there was a time when we hesitated to leave our houses, waiting for the situation to settle down a bit.”* (Female CHW, FDG, district hospital)

CHWs reported a decline in community referrals and contact tracing, as clients feared contracting COVID-19 at health facilities and unwilling COVID-19 vaccination:

> *“…you can reach the place you are going [and] they may not cooperate because when they see that container [cooler box], they think that they are going to be vaccinated [for COVID-19] against their will…”* (Female CHW, FGD, health center)
>
> *“…after you get a suspect and you manage to bring him/her to the hospital for medication, you must go back and test the children and other household members. They refused to take their children to the hospital for fear that they would contract COVID-19.”* (Female CHW, FGD, district hospital)

Some CHWs stated that communities were reluctant to welcome them during home visits, often providing false addresses or names due to fear of COVID-19 exposure. One CHW reported that some people were unwilling to provide sputum samples despite showing TB symptoms:

> *“…you can find someone; when you talk to him, he tells you, “I don’t have sputum. I cannot produce sputum, sir”, and you see him/her coughing. [You tell him] to provide [sputum, but he insists] “Sir, I am not coughing…”* (Male CHW, IDI, district hospital)

### Adaptations for sustaining TB ACF during COVID-19

CHWs perceived themselves at high risk for contracting COVID-19 while conducting TB ACF. However, they continued implementing ACF activities with COVID-19 precautions, such as wearing masks and gloves, sanitizers, and social distancing. They prioritized persons at most risk of contracting COVID-19 i.e., those living with HIV, hypertension, and diabetes:

> *“We gave priorities to most of the persons living with HIV/AIDS…high blood pressure, diabetes… and this was after being trained that most studies show that people with diabetes…are at high risk of contracting Tuberculosis and… more at risk to contract COVID-19…”* (Male CHW, IDI, regional hospital)

Some CHWs mentioned using phone calls to contact clients instead of making home visits and requested treatment supporters to collect drugs for their clients.

> *“…we mostly depended on calling [TB clients] either to invite them to the clinic or their treatment supporters to collect [TB drugs] on their behalf… so they did not stop taking the medication; instead, they reduced their frequency of coming to the clinic…”* (Male CHW, IDI, health center)

## DISCUSSION

This study describes difficulties caused by COVID-19 in supplying TB commodities and maintaining TB care services in Tanzania during the first year of the pandemic. We observed stockouts of GeneXpert cartridges, sputum containers, and pediatric TB medications alongside disruptions in TB care processes, resulting in decreased TB screening and contact tracing. Healthcare providers adapted by using alternative containers for sputum collection, different TB diagnosis platforms, tailored differentiated services for TB risk groups, extended TB drug refill schedules, and used mobile communication for contact tracing.

We observed shortages in pediatric TB drugs but not in adult TB drugs, indicating that the pandemic’s impact on TB commodities varied, unlike in other high TB burden countries that reported TB drug shortages for both children and adults^24^. The redistribution of commodities among facilities may have ensured the availability of adult TB drugs. However, stockouts of pediatric TB drugs align with other studies on TB supply chain during the pandemic^25^. Possible causes include donor funding dependency, which unfortunately shifted to COVID-19 prevention efforts, international transport issues due to lockdown policies, low manufacturing capacity, and increased commodity costs^26^. TB commodity stockouts, particularly for GeneXpert cartridges and pediatric TB drugs, have also been reported during pre-pandemic times in Tanzania^16^ and other countries within the WHO African region,^27^ and is argued to be a foremost challenge in ending TB by 2030. Therefore, targeted efforts are needed to strengthen the supply chain post-pandemic.

The decline in utilization of TB services was also experienced globally at varying magnitudes and often attributed to fear and stigma of COVID-19^28,29^, similar to the underlying reasons in Tanzania. Taking account of this impact, together with increased TAT from overreliance on microscopy and/or chest X-ray, these may have influenced the country’s minimal declines in TB case notifications^16,25^, particularly during the first year of the pandemic. Nevertheless, unlike Tanzania, other low- and middle-income countries (LMICs) encountered additional barriers to utilizing TB services, namely: lockdowns, transport disruptions, and increased transportation costs that hindered the movements of health providers and clients to health facilities^30^.

Our findings enhance our understanding of how TB services were maintained in Tanzania during the COVID-19 pandemic. The mitigations taken clarify the country’s ability to lessen the impact of COVID-19 on TB service delivery, thus enabling an understanding of the minimal fluctuations in TB case notifications reported in published literature^17,24,25^. The adaptations made in TB clinical practices provide valuable insights for future pandemic preparedness and efforts to strengthen TB control^31^. National Tuberculosis Control Programs could adopt some of these adaptations post-pandemic. For instance, the resort to home-based care, tracing by mobile communication, and extended drug refill schedules is similar to other countries^30,32–35^ and can be extended for TB treatment post-pandemic. The role of digital technology in supporting TB services delivery during the pandemic is documented by varied case studies compiled by the WHO during the first year of the pandemic^36^ and recommended as a catch-up strategy to mitigate the impact of COVID-19 on TB services^37^. This informs further research on the multi-month dispensing of TB drugs coupled with teleconsultations and telemonitoring for TB care. The challenges and service adaptations are lessons learned for future TB service delivery beyond pandemics (Table 4).

### Lessons from TB service delivery during COVID-19

|  | **Challenges encountered** | **Adaptations** | **Lessons for TB service delivery** |
| --- | --- | --- | --- |
| 1. | Stockouts and delays in TB commodities | -Use of alternative sputum sample containers<br>-Sharing commodities among facilities<br>-GeneXpert prioritization for TB Key and vulnerable populations<br>-Use of sputum pooling system<br>-Adjustment of adult TB drugs for pediatrics | -Strengthen the TB supply chain for resilience during emergencies<br>-Establish networks for sharing TB commodities and real-time communication<br>-Integration of various diagnostic tools using different samples |
| 2. | Hesitancy to seek TB services due to fear of contracting or diagnosis with COVID-19 with subsequent quarantine | -Alternative drug dispensing (home delivery by CHWs, facility gate pick-ups)<br>-Extended drug refill schedules (biweekly for intensive and monthly for continuation)<br>-Mobile phone follow-ups | -Adoption of varied TB drug dispensing strategies that allow remote access for clients<br>-Research multi-month drug refill schedules for stable persons with TB<br>-Utilize mHealth technology to enhance service delivery<br>-Inform communities about infection control measures in health facilities |
| 3. | PPE shortages led HCWs to use fabric masks or wear masks longer, increasing<br>infection risks | HCWs reused PPE, purchased fabric masks out-of-pocket, and sourced PPE from donors | -Sustain HCW protection in emergencies |

The first year of the pandemic coincided with the reign of the former late President Dr. John Pombe Magufuli, who did not impose total lockdown and emphasized sustained service delivery^38,39^. This enabled HCWs and clients to access health facilities without travel restrictions, facilitating the redistribution of TB commodities and adaptations in TB clinical practices for sustained service delivery.

This qualitative study cannot quantify the impact of reported challenges on TB services. We did not explore experiences throughout the entire pandemic period; instead, we focused on retrospective insights into the pandemic’s impact on TB during its peak, when most resources shifted to COVID-19. While these retrospective accounts may affect the accuracy of reported challenges and adaptations, triangulating data collection methods helped mitigate recall bias by allowing participants to collectively recall and validate their experiences. Additionally, focusing on high-COVID-19 regions and TB facilities that provide COVID-19 care may limit the transferability of findings to areas with lower COVID-19 burdens, where TB service delivery challenges may differ. Nevertheless, our findings provide insights into healthcare providers’ TB management experiences during the first two COVID-19 waves.

## CONCLUSION

COVID-19 disrupted TB commodity availability and TB treatment-seeking behavior. Adaptations like multi-month drug refills and optimized GeneXpert cartridge use supported the TB healthcare system’s resilience. While these adaptations offer valuable insights for strengthening TB service delivery, their effectiveness and sustainability require further evaluation. Thus, prospective studies could clarify their long-term impact. National Tuberculosis Programs could consider adapting these practices post-pandemic, with appropriate modifications to suit different contexts.

## Supporting information

Supplemental data collection tools

## Data availability statement

The data that supports the study findings are available upon reasonable request from the corresponding author (DP).

## Acknowledgements

We are grateful to the participants who consented to participate in the study and acknowledge the contributions of research assistants in supporting data collection.

## Contributors

DP: study conceptualization, data collection and analysis, wrote the first draft of the manuscript, critical review and editing. WW: study conceptualization, critical review and editing. LM: study conceptualization, data collection and analysis, critical review and editing. ES: data collection and analysis, wrote the first draft of the manuscript, critical review and editing. HM: data collection and analysis, wrote the first draft of the manuscript, critical review and editing. HO: data collection and analysis, wrote the first draft of the manuscript, critical review and editing. TS: wrote the first draft of the manuscript, critical review and editing. CM, WO, IS, RB, EM, RK, AT, LM, NEN: critical review and editing.

## Supplementary materials

i. S1: Study protocol
ii. S2: Topic guide for HCWs
iii. S3: Topic guide for CHWs
iv. S4: In-depth interview guide for HCWs
v. S5: In-depth interview guide for CHWs
vi. COREQ checklist

