## Supplemental data collection tools for "Tuberculosis services delivery challenges and their mitigations during the COVID-19 pandemic in Tanzania: A qualitative study"

### IN-DEPTH INTERVIEW GUIDE (Health workers)

**Interview Information**
Name of Interviewer:
Interviewee code:
Date:
Duration of interview:

**Introductory Remarks:**

Thank you for consenting to participate in this in-depth interview. I am......................... and I would like to explore your views about the impact of COVID-19 on the quality of TB healthcare services provision and uptake during the first year of the COVID-19 pandemic.

### PART A: DEMOGRAPHICS

Facility name:

Sex:

Profession:

Position/role:

Experience in provision of TB treatment (years):

### PART B: WORKPLACE EXPOSURE TO COVID-19

1. To begin with, I would first like to understand how you prevented yourself against exposure to COVID-19 from the time when it was announced that the country had a COVID-19 case.
    - types of PPE used-how were they financed?
    - prevention during shortage of PPEs

2. How did you perceive your risk for COVID-19 exposure in the first year that the country was affected?
    - low/high risk
    - why? interaction with TB patients, use of PPEs

3. How did persons with TB perceive their risk for COVID-19 when attending treatment at the facility?

- concerns raised, use of PPEs

4. If you think about the times when you had suspected or confirmed COVID 19 cases in your facility, how did their treatment affect TB service delivery at the facility?
    - changes in screening, diagnosis and treatment of TB

5. How did you differentiate between COVID 19 and presumptive TB during the COVID 19 period?
    - source of knowledge
    - confirmation from test results

### PART C: PROCUREMENT AND DISTRIBUTION OF TB COMMODITIES

1. What changes did you observe in funding for TB commodities in the first year that the country was affected by COVID-19?
    - drugs, cartridges, reagents, masks, sputum containers
    - shortage of funding
    - reallocation of TB funding to COVID-19 PPEs and care
    - additional funding from donors

2. What changes did you observe in procurement and distribution of TB commodities in the first year that the country was affected by COVID-19?
    - drugs, cartridges, reagents, masks, sputum containers
    - time to receipt of orders
    - changes of order quantity
    - out of stocks

3. How long did the observed changes in question 2 last?
    - why that long?

4. How did you adapt to changes in availability of TB commodities in the first year that the country was affected by COVID-19?
    - requesting nearby facilities/countries
    - demand forecasting
    - inventory management strategies
    - drug suppliers with short lead times

### PART D: TB SCREENING, DIAGNOSIS AND TREATMENT

1. How were clients screened for TB in the first year that the country was affected by COVID-19?
    - limits to daily screening
    - changes in client facility utilization
    - changes in CHW referrals
    - availability of staff
    - Microscopy, GeneXpert, culture-why?

2. What challenges did you experience in provision of TB screening services during the COVID-19 period?
    - fear of COVID-19
    - reduced community referrals

3. What measures were taken to ensure sustained provision of TB screening services at the facility during COVID 19 period?
    - Use of PPEs
    - limits to daily screening
    - contacting CHWs

4. What challenges did you experience in diagnosis of TB during the first year that the country was affected by COVID-19?
    - availability of cartridges, drugs, reagents, sputum containers, power cuts

5. How were TB presumptive cases diagnosed in the shortage of cartridges and reagents during the first year that the country was affected by COVID-19?
    - Sputum pooling system
    - MDR TB diagnosis

6. What changes in the specimen referral system did you observe during the COVID-19 period?
    - handling of sample referrals
    - changes in TAT

7. How was TB contact tracing performed during the COVID-19 period?
    - role of CHWs
    - challenges experienced

8. How was TB treatment managed during the COVID-19 period?
    - limits to daily no. of clients
    - prioritization of patients based on certain criteria

9. How were patients categorized as lost to follow up managed during the COVID 19 period?
    - tracing activities

10. What impact does COVID-19 vaccination have on TB service delivery during this COVID-19 pandemic?
    - screening activities
    - diagnosis
    - treatment
    - patient perspectives about COVID-19 vaccine

11. What recommendations do you have on ways to sustain delivery of TB services during this COVID-19 pandemic?
    - activities to support screening, diagnosis and treatment

**IN-DEPTH INTERVIEW GUIDE (Community Health Workers)**

**<u>Interview Information</u>**
Name of Interviewer:
Interviewee code:
Date:
Duration of interview:

**Introductory Remarks:**

Thank you for consenting to participate in this in-depth interview. I am......................... and I would like to explore your views about the impact of COVID-19 on the quality of TB healthcare services provision and uptake during the first year of the COVID-19 pandemic.

### PART A: DEMOGRAPHICS

Name of facility supported by CHW:

Sex:

Educational level:

Experience in TB active case finding (years):

### PART B: EXPOSURE TO COVID-19

1. To begin with, I would first like to understand how you prevented yourself against exposure to COVID-19 from the time when it was announced that the country had a COVID-19 case.
    - types of PPE used-how were they financed?
    - prevention during shortage of PPEs
    - challenges experienced
    - source of prevention knowledge

2. How did you perceive your risk for COVID-19 exposure in the first year that the country was affected?
    - low/high risk
    - why? interaction with TB patients, use of PPEs

3. How did people presumed to have TB perceive their risk for COVID-19?
    - concerns raised
    - high/low risk-why?

4. How did you differentiate between COVID 19 and presumptive TB during the COVID 19 period?
    - source of knowledge
    - confirmation from results

**PART C: TB ACTIVE CASE FINDING ACTIVITIES**

1. How were ACF activities structured in pre-COVID-19 period?
    - challenges experienced
    - awareness campaigns conducted in communities

2. What changes were made in the implementation of ACF during COVID-19 period?
    - changes in screening in communities
    - changes in community referrals
    - limits to daily screening at the facility
    - prioritization of target groups

3. What challenges did you experience in the conduct of ACF during COVID-19 period?
    - shortage/increased support from donors?
    - fear to visit facilities (clients and CHWs)
    - delays in diagnosis and TAT for receipt of results

4. What measures were taken to ensure sustained identification of presumptive TB cases in communities during COVID 19 period?
    - use of PPEs
    - limits to daily screening

5. How was TB contact tracing performed during the COVID-19 period?
    - challenges experienced
    - facility support

6. How was support for TB treatment provided during the COVID-19 period?
    - limits to daily no. of clients

- prioritization of patients based on certain criteria
- changes compared to pre-COVID 19 period

7. How was support for patients categorized as lost to follow up managed during the COVID 19 period?
    - tracing activities

8. What effects does COVID-19 vaccination have on TB ACF activities during this COVID-19 pandemic?
    - community screening activities
    - contact tracing
    - tracing LTFUs

**TOPIC GUIDE (Health workers)**

**Focus group discussion Information**

Name of facilitator:

FGD code:

Date:

Duration of discussion:

### PART B: WORKPLACE EXPOSURE TO COVID-19

1. To begin with, we would first like to understand how you prevented yourself against exposure to COVID-19 from the time when it was announced that the country had a COVID-19 case.
    - types of PPE used-how were they financed?
    - prevention during shortage of PPEs
    - perception of level of risk-why?

2. How did persons with TB perceive their risk for COVID-19 when attending treatment at the facility?
    - concerns raised, use of PPEs

3. How did you differentiate between COVID 19 and presumptive TB during the COVID 19 period?
    - source of knowledge
    - confirmation from test results

### PART C: PROCUREMENT AND DISTRIBUTION OF TB COMMODITIES

1. What changes did you observe in procurement and distribution of TB commodities in the first year that the country was affected by COVID-19?
    - Changes in funding- reallocation to COVID-19, additional funding
    - drugs, cartridges, reagents, masks, sputum containers
    - time to receipt of orders
    - changes of order quantity
    - out of stocks
    - how long did they last?

2. How did your facility adapt to changes in availability of TB commodities in the first year that the country was affected by COVID-19?
    - requesting nearby facilities/countries
    - demand forecasting
    - inventory management strategies
    - drug suppliers with short lead times

### PART D: TB SCREENING, DIAGNOSIS AND TREATMENT

1. How was screening, diagnosis and treatment for TB conducted in the first year that the country was affected by COVID-19?
    - limits to daily screening
    - changes in client facility utilization
    - changes in CHW referrals
    - availability of staff
    - reduced community referrals
    - microscopy, GeneXpert, culture-why?
    - challenges experienced-cartridges, drugs, reagents, sputum containers
    - diagnosis in times of shortage of TB commodities (sputum pooling system)

2. What challenges did you experience in screening, diagnosis and treatment for TB during the COVID-19 period?
    - fear of COVID-19

3. What measures were taken to ensure sustained provision of TB screening, diagnosis and treatment services at the facility during COVID 19 period?
    - use of PPEs
    - limits to daily screening
    - contacting CHWs

4. What changes in the specimen referral system did you observe during the COVID-19 period?
    - handling of sample referrals
    - changes in TAT

5. What impact does COVID-19 vaccination have on TB service delivery during this COVID-19 pandemic?
    - screening activities
    - diagnosis
    - treatment
    - patient perspectives about COVID-19 vaccine

6. What recommendations do you have on ways to sustain delivery of TB services during this COVID-19 pandemic?
    - activities to support screening, diagnosis and treatment

**TOPIC GUIDE (Community Health Workers)**

**Focus group discussion Information**
Name of facilitator:
Discussion code:
Date:
Duration of discussion:

### PART B: EXPOSURE TO COVID-19

1. To begin with, I would first like to understand how you prevented yourself against exposure to COVID-19 from the time when it was announced that the country had a COVID-19 case.
    - types of PPE used-how were they financed?
    - prevention during shortage of PPEs
    - challenges experienced
    - source of prevention knowledge

2. How did you perceive your risk for COVID-19 exposure in the first year that the country was affected?
    - low/high risk
    - why? interaction with TB patients, use of PPEs

3. How did people presumed to have TB perceive their risk for COVID-19?
    - concerns raised
    - high/low risk-why?

4. How did you differentiate between COVID 19 and presumptive TB during the COVID 19 period?
    - source of knowledge
    - confirmation from results

### PART C: TB ACTIVE CASE FINDING ACTIVITIES

1. How were ACF activities structured in pre-COVID-19 period?
    - challenges experienced

2. What changes were made in the implementation of ACF during COVID-19 period?

- changes in screening in communities
- changes in community referrals
- limits to daily screening at the facility
- prioritization of target groups

3. What challenges did you experience in the conduct of ACF during COVID-19 period?
    - shortage/increased support from donors?
    - fear to visit facilities (clients and CHWs)
    - delays in diagnosis and TAT for receipt of results

4. What measures were taken to ensure sustained identification of presumptive TB cases in communities during COVID 19 period?
    - use of PPEs
    - limits to daily screening

5. How was TB contact tracing performed during the COVID-19 period?
    - challenges experienced
    - facility support

6. How was support for TB treatment provided during the COVID-19 period?
    - limits to daily no. of clients
    - prioritization of patients based on certain criteria
    - changes compared to pre-COVID 19 period

7. How was support for patients categorized as lost to follow up managed during the COVID 19 period?
    - tracing activities
